# Incidence of SARS-CoV-2 infection among asymptomatic frontline health workers in Los Angeles County, California

**DOI:** 10.1101/2020.11.18.20234211

**Authors:** Megan Halbrook, Adva Gadoth, Rachel Martin-Blais, Ashley Grey, Deisy Contreras, Saman Kashani, Clayton Kazan, Brian Kane, Nicole H. Tobin, Jennifer A. Fulcher, Sarah L. Brooker, Scott G. Kitchen, Emmanuelle Faure-Kumar, Otto O. Yang, Kathie G. Ferbas, Grace M. Aldrovandi, Anne W. Rimoin

## Abstract

Beginning April 8, 2020, we enrolled 1787 frontline heath workers who were asymptomatic for COVID-19 into a longitudinal surveillance study. During that time 4 healthcare workers and 6 first responders tested positive for SARS-CoV-2 by RT-PCR. Additionally, 43 healthcare workers and 55 first responders had detectable IgG antibodies to SARS-CoV-2.

## Introduction

Our understanding of the epidemiology and pathogenesis of severe acute respiratory syndrome coronavirus 2 (SARS-CoV-2) infection remains incomplete. Frontline health workers are at heightened risk for exposure to and transmission of SARS-CoV-2 due to their close physical contact with persons requiring medical attention and interventions and the close work and rest spaces. They also represent an ideal population for the study of asymptomatic and minimally symptomatic spread of COVID-19, with vested interests in frequent screening [1, 2]. With this in mind, we enrolled a cohort of asymptomatic persons employed by the University of California, Los Angeles (UCLA) Health system and the Los Angeles County Fire Department (LACoFD) into a longitudinal research study designed to assess SARS-CoV-2 attack rates, exposure risks, and correlates of immunity during the course of the COVID-19 pandemic in Los Angeles County.

## Methods

UCLA Health employs over 38,000 individuals across four hospitals and over 180 primary care practices in Los Angeles County. LACoFD is responsible for the emergency response of 4 million residents across Los Angeles County, and staffs 175 fire stations and all public lifeguard towers with approximately 4000 frontline personnel. Beginning on April 8, 2020 and May 19, 2020, respectively, health system workers (HSW) employed by UCLA Health—including those with and without direct patient care responsibilities— and LACoFD first responders (FR) from eight battalions— including firefighters, paramedics, lifeguards, and dispatchers—were invited to enroll in this study. Eligible participants were over 18 years of age and free of new symptoms associated with COVID-19 in the 2 days prior to enrollment.

Participants were asked to provide monthly blood samples and up to biweekly self-collected mid-turbinate nasal swabs at their workplace; participants also responded to online questionnaires following sample collection to assess demographics and potential exposures to SARS-CoV-2. Nasal swabs were tested for the presence of SARS-CoV-2 using the FDA EUA approved Abbott Molecular dual target real-time multiplex PCR assay for the detection of SARS-CoV-2 RNA. Blood samples were analyzed by enzyme-linked immunosorbent assay (ELISA) to detect anti–SARS-CoV-2 spike receptor-binding domain IgG, as previously described [3, 4].

Tabular counts, attack rates and associations between seroprevalence and demographic factors and occupational exposures were calculated using SAS statistical software version 9.4 (Cary, NC). While some participants may have received additional outside testing prior to enrollment or in between study visits, test results shown here reflect only SARS-CoV-2 PCR and serology performed as a part of study activities.

This study was approved by the UCLA institutional review board on March 25, 2020. New enrollments and follow-up visits are ongoing; results reported here reflect data gathered through August 31, 2020.

## Results

From April through August, 2020, we enrolled 1108 HSW and 679 FR. Those aged 60 and older accounted for 4.9% of HSW and 2.2% of FR. Women accounted for almost two-thirds of enrolled HSW (64%), but only a small minority of study FR (6.0%). A majority (add % that include at least 1 exposure) of frontline workers in both groups reported conducting procedures with potential for exposure to COVID-19 patients, clinical samples, or work spaces (Table 1).

**Table 1.**
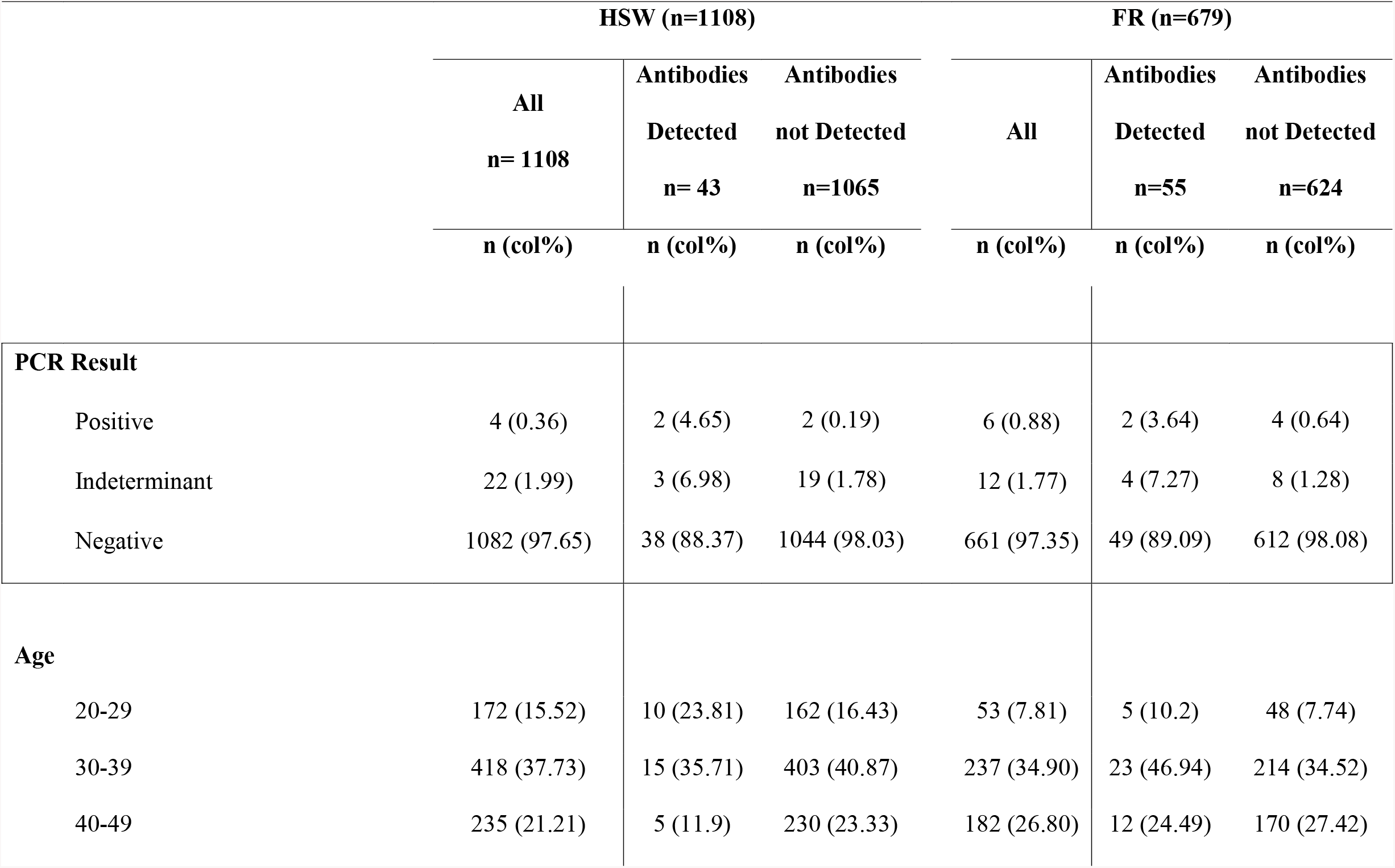
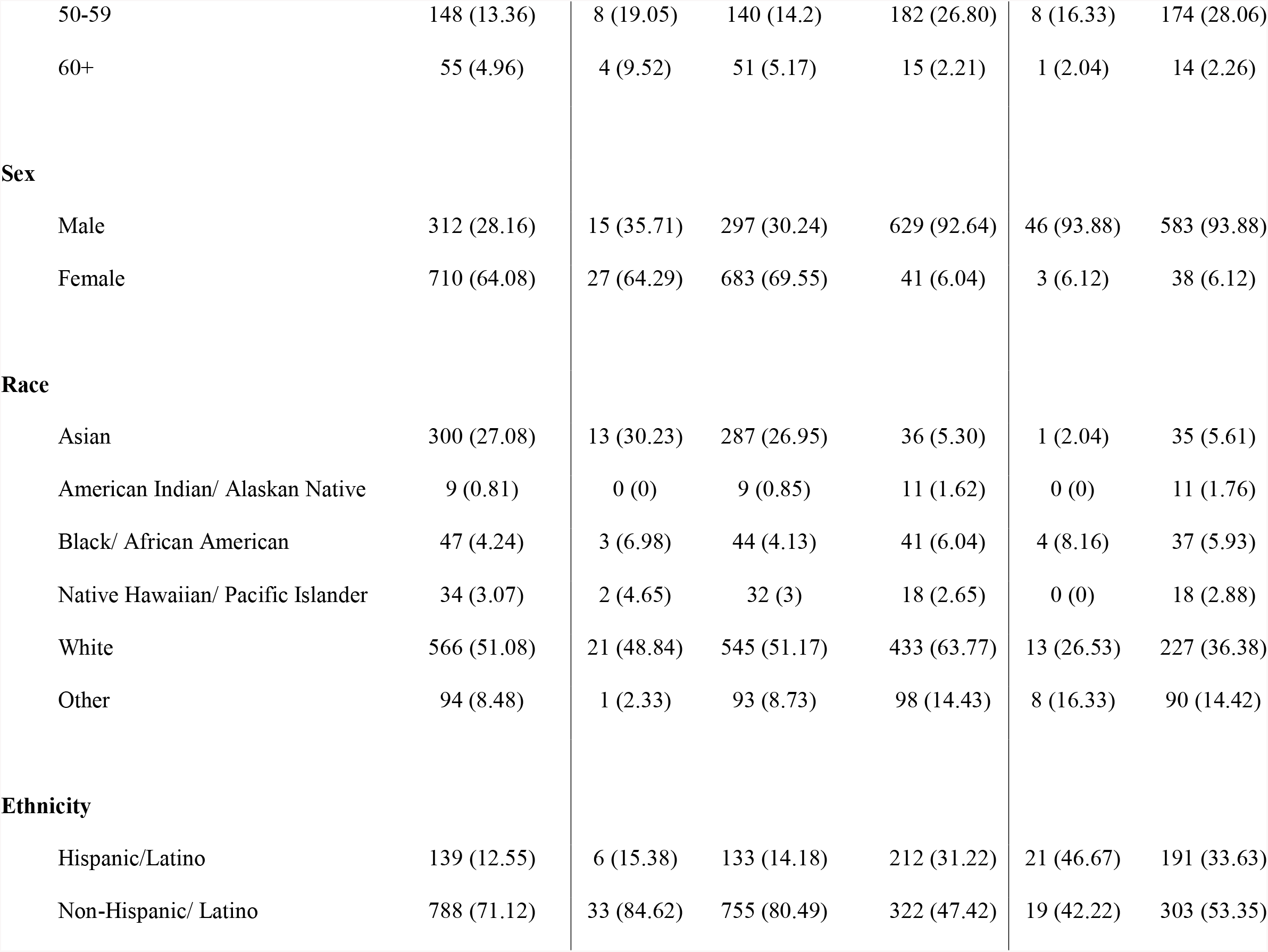
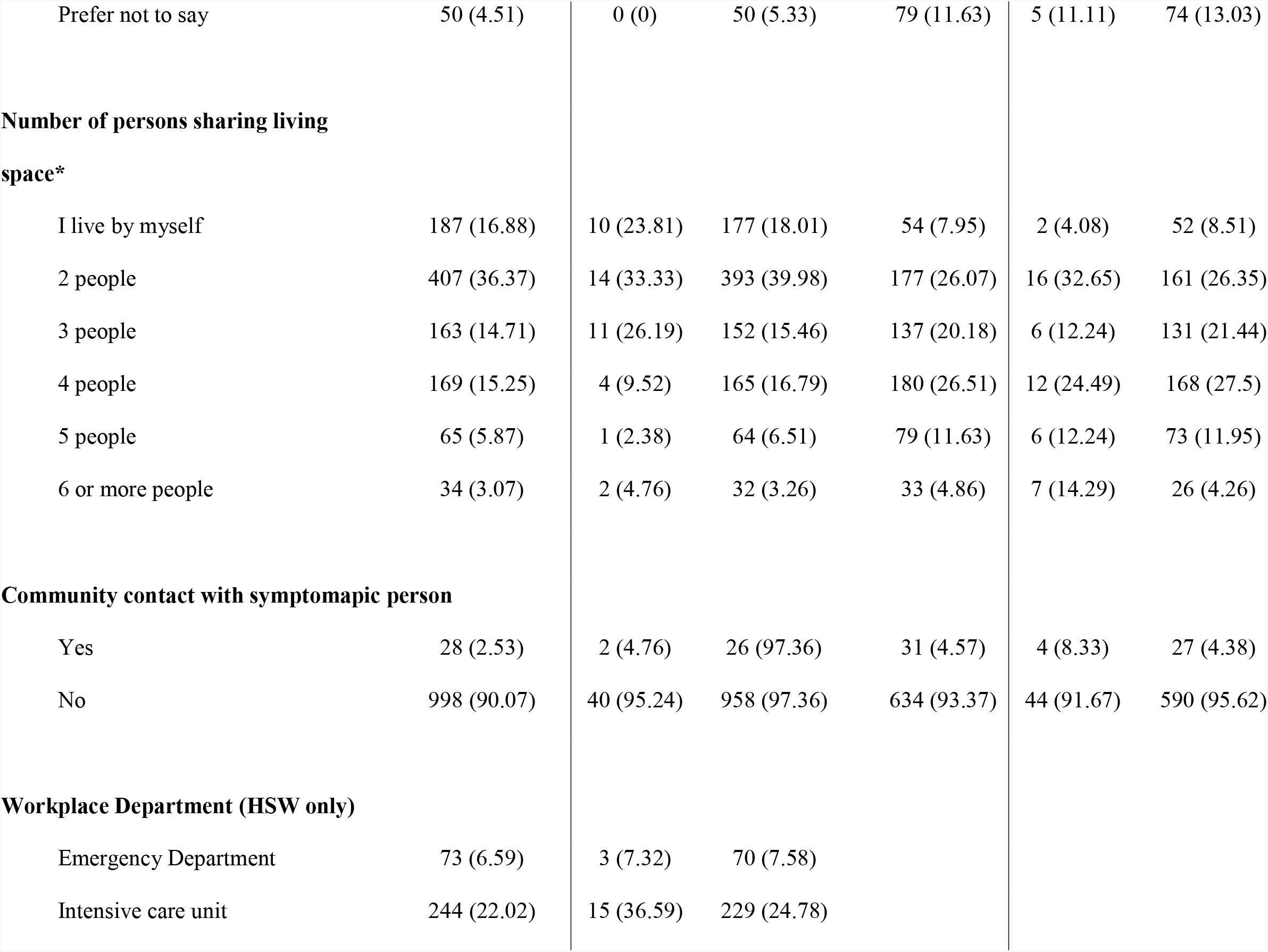
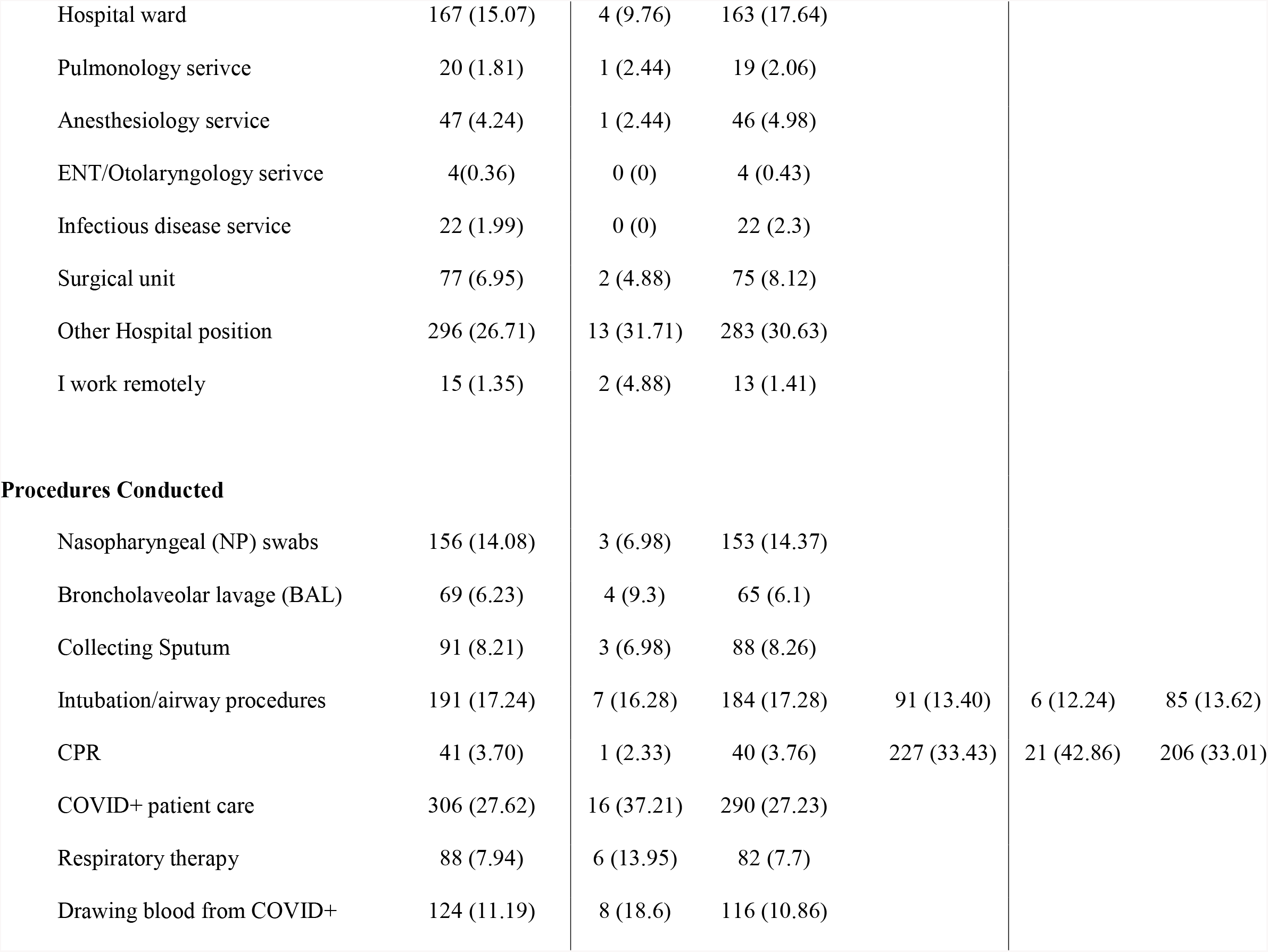
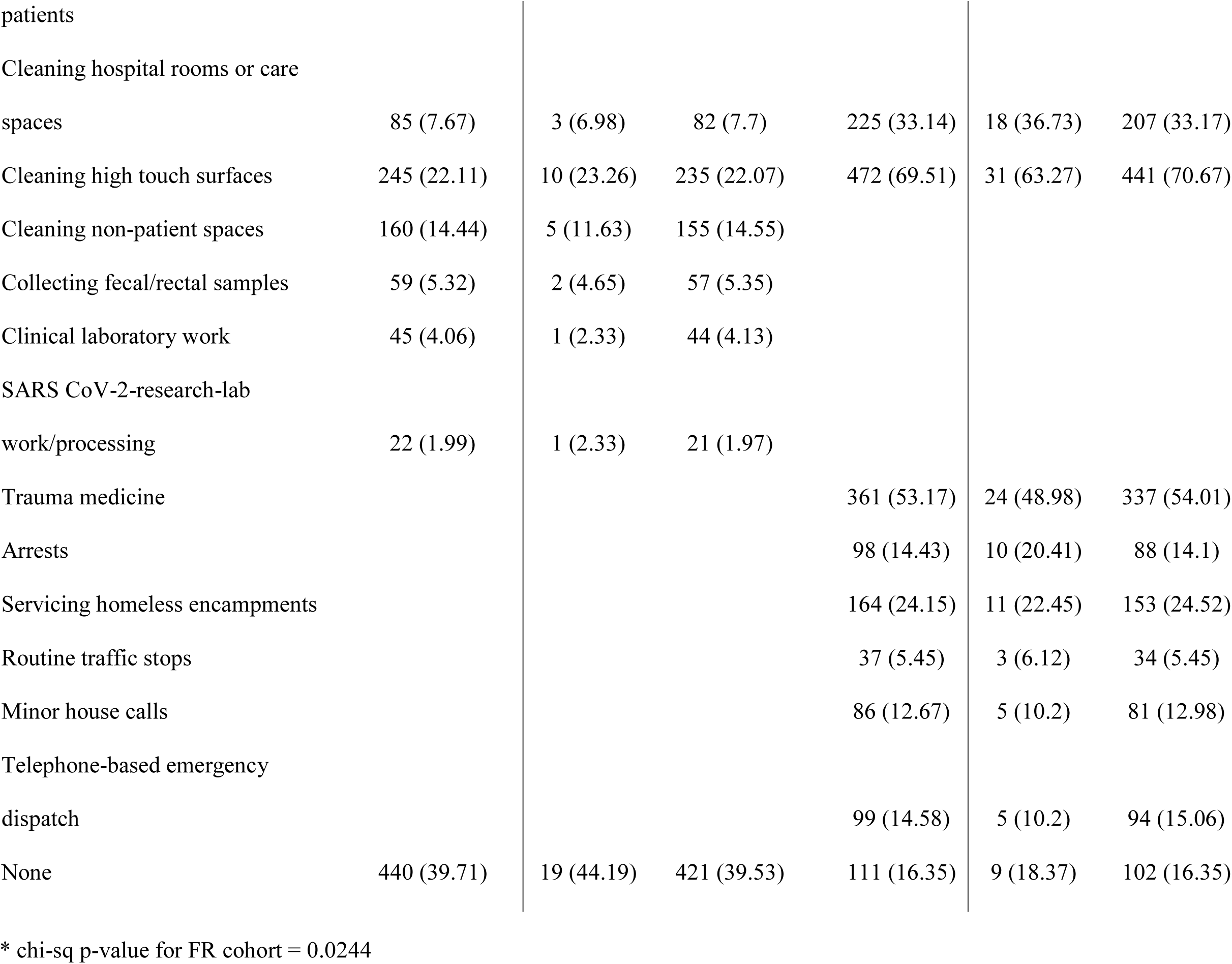
Test Results & Participant Demographics.

During these initial months of observation, 4 (0.4%) HSW and 6 (0.9%) FR tested positive for SARS-CoV-2 by RT-PCR, with 22 (2%) HSW and 12 (1.8%) FR returning indeterminate PCR results. Thus, a five-month attack rate of 0.37 – 2.4% among HSW, and a four-month attack rate of 0.91 – 2.72% was determined for FR. Attack rates between these two groups were not significantly different from one another.

Forty-three HSW (3.9%) had or developed detectable IgG antibodies to SARS-CoV-2 during the study period; among FR, 55 (8.1%) had detectable IgG. Among those with indeterminate PCR tests, 19 (86%) HCW and 8 (66%) first responders did not have or had not yet developed detectable antibodies. Thirty-eight (3.4%) HSW and 49 (7.2%) FR had detectable SARS-CoV-2 antibodies without known history of infection during the initial study period (Table 1). Notably, among those who tested positive by PCR, two-thirds of first responders (n= 4) and half of HSW (n= 2) displayed active infection in late June.

Across both cohorts, rates of seroreactivity did not differ by demographic characteristics, job role, or performed medical procedures. Among the FR, sharing your living space with six or more people was associated with detection of SARS-CoV-2 antibodies (p=0.024). The odds of seroreactivity among those living with six or more persons was 7 times greater compared to those living alone (95% CI: 1.36 -36.10). This relationship was not observed across other household sizes (2-5 persons), nor among the HSW.

## Discussion

These preliminary baseline data represent the first descriptions of an ongoing longitudinal study of frontline workers in Los Angeles county. As a whole, rates of active infection in this population were lower than in the general population as reported by the LA County Department of Public Health (LACDPH) during the same period [5]. However, a late June peak in active infections among our cohort does appear to mirror the larger LA community, which saw its positive test rate spike in mid-July.

The low overall incidence observed could be indicative of proper and widespread use of personal protective equipment in these high-risk occupational settings, as universal masking protocols were in place in both work settings. Healthy worker bias is likely also an important explanatory factor in our recruitment of asymptomatic and mostly younger adults (below age 60), and in our finding of low infection prevalence among a population at the front lines of SARS-CoV-2 exposure.

Our results are contextualized by SARS-CoV-2 infection and exposure rates demonstrated in other studies of frontline health workers. In a convenience sample from 13 academic medical centers across the United States (including UCLA Health), only 6% of healthcare workers caring for COVID-19 patients displayed antibody evidence of SARS-CoV-2 infection [6]. Other analogous US health worker cohorts found rates of PCR positivity between 3.6-6.5% and antibody prevalence of 7.6% [7, 8]. The lower prevalence of serologic markers for SARS-CoV-2 infection observed in our study makes sense given our inclusion of workers across a spectrum of direct, indirect, and no occupational exposure to COVID-19 patients.

An investigation of SARS-CoV-2 infections by the LACDPH found that, from February-May, 2020, 10% of incident cases reported across the county occurred among healthcare workers [9]. Thus, despite low rates of incident infection observed among HSW and FR, frontline health workers make up an important proportion of new cases identified across the county. Notably, LACDPH investigated positive-testing healthcare workers across 27 distinct settings and identified correctional facilities, schools, and long-term care facilities as outbreak hotspots [9]. In contrast, this study enrolled workers from hospital, outpatient, and first response settings, which may help account for the lower prevalence of positive PCR tests and antibody markers observed. While some high-risk vocations weren’t targeted by this study, 11% of HSW reported secondary healthcare jobs in these settings: fifteen participants held positions at long-term care facilities and two reported additional roles at colleges.

While this study was open to all LACoFD and UCLA Health employees, work schedules and study logistics may have created barriers to participation. Rolling enrollment from April 8 to August 31, 2020 may also have introduced some measure of temporal confounding into our analysis, given the variable testing infrastructure, governmental policies, and communal incidence of SARS-CoV-2 infection in LA County over this timeframe. Thus, further analysis at the conclusion of study activities should include an enrollment date-adjusted examination of biological markers of infection.

## Data Availability

Data is not publicly available but interested parties should contact study authors for inquiries regarding data access.

## Acknowledgements

The authors would like to acknowledge the contributions of Patrick Arena, Angie Barrall, Cindy Beard, Sylvia Tangney, Michael Mengual, Marjorie Weiman, Gabby Merlo, Sergio Duron, Faith Landsman, Sarah Zabih, Ana Zamora, Monica Saavedra, Hwee Ng, Lorena Porras-Javier, and La Quinta N. Montgomery.

## Footnote

This work was supported by AIDS Healthcare Foundation, The Shurl and Kay Curci Foundation, Elizabeth R. Koch Foundation, The Horn Foundation, and Steven & Alexandra Cohen Foundation.

The authors have no conflicts of interest to declare.

